# Mental Health Status of the General Population during the COVID-19 Pandemic: A Cross-Sectional National Survey in Japan

**DOI:** 10.1101/2020.04.28.20082453

**Authors:** Michiko Ueda, Andrew Stickley, Hajime Sueki, Tetsuya Matsubayashi

## Abstract

**Aims:** The ongoing COVID-19 pandemic may have detrimental mental health consequences. However, there is limited understanding of its impact on the mental health of the general population. The aim of this study was to examine the mental health of the Japanese general population by conducting the first systematic survey during the pandemic with a particular focus on identifying the most vulnerable groups. Methods: Data was obtained from an online commercial web panel of 2000 respondents in April and May 2020. Information was collected on demographic and socioeconomic factors as well as mental health status (anxiety and depressive symptoms). Logistic regression analysis was used to examine associations.

**Results:** The mental health of young and middle-aged individuals was significantly worse than that of older individuals during the pandemic. There was also some indication that individuals who were not currently working were significantly more likely to report a high level of anxiety and depressive symptoms. Part-time and temporary contract-based workers were also more likely to suffer from depressive symptoms.

**Conclusion:** Our results highlight that monitoring the mental health of younger and economically vulnerable individuals may be especially important. In addition, they also indicate that population mental health might not only be affected by the direct health consequences of COVID-19, but also by the economic ramifications of the pandemic.

## 1. Introduction

In late December 2019, the novel coronavirus (COVID-19) was detected in the city of Wuhan, China. Within weeks, rapid person-to-person transmission resulted in widespread infection inside Wuhan.^1^ National and international travel subsequently facilitated the swift spread of the virus to Europe, North America, Asia, and the Middle East, so much so, that there were more than 118,000 cases in 114 countries when the World Health Organization (WHO) characterized the COVID-19 situation as a pandemic on March 11, 2020.^2,3^ The rapid dissemination of the virus has also been accompanied by a large number of deaths in countries across the world, with individuals who are older and who have multiple comorbidities seemingly especially vulnerable to the disease.^4,5^

In the absence of a vaccine, and given the high degree of transmissibility and potential lethality of COVID-19, social and physical distancing, including reducing/avoiding crowding, the closure of non-essential businesses/services, stay-at-home orders, protecting vulnerable groups and local/national movement restrictions, have been the main public health measures adopted to mitigate the transmission/detrimental effects of the virus.^6^ Despite the potential benefits of such measures, it is also being increasingly recognized that they might also have both negative short- and long-term consequences for mental health^7^ that are in addition to those resulting from the disease itself.^8^ For example, a recent review article has shown that quarantine may be associated with a number of negative psychological outcomes, such as anger and posttraumatic stress, with factors such as financial loss and the socioeconomic distress that can result from quarantine possibly underpinning the emergence of psychological disorders.^9^

Several factors suggest that a focus on mental health might be especially important in relation to the ongoing COVID-19 pandemic. First, studies on earlier global infectious disease outbreaks, such as the severe acute respiratory syndrome (SARS) epidemic (2002-2004) and the Middle East respiratory syndrome occurrence (MERS; 2012-), have indicated that the spread of infectious disease is linked to worse mental health within and across populations.^10,11^ Second, recent studies from China have similarly shown that COVID-19 may be important for mental health outcomes such as anxiety, depression, and posttraumatic stress symptoms.^12,13,14^ Third, it is possible that COVID-19 may disproportionately affect vulnerable populations,^15^ with groups such as older adults, those with co-occurring illnesses, the socially excluded (e.g., the homeless), and individuals with low incomes all being at an increased risk for poorer mental health.^16^ Given the lack of evidence, determining who is vulnerable to the mental health consequences of COVID-19 is essential for the design and implementation of evidence-based interventions to mitigate the potential effects of COVID-19 on the population’s mental health.

This study will examine the effects of the COVID-19 pandemic on the mental health of a national sample of the Japanese population. The first case of COVID-19 was registered in Japan on January 16, 2020 in the Kanagawa prefecture.^17^ Cases continued to grow against a backdrop of increasingly strong public health interventions including the temporary closure of schools and postponement of the forthcoming Olympics in March.^18^ A sudden spike in the number of cases led to the declaration of a state of emergency on April 7 in seven major prefectures, including Tokyo.^18^ Following an upsurge in ‘untraceable’ new infections, the state of emergency was expanded to cover the whole of Japan, and thirteen prefectures were designated as “special alert areas” on April 16.^19^ Although there is no comprehensive data yet, there is some indication that these COVID-19-related events may be having a negative impact on population wellbeing in Japan. For example, the prospect of a possible forthcoming lockdown resulted in panic buying of food and other household products in Tokyo at the end of March^20^, while a recent international poll showed that 86% of the Japanese population were afraid of catching the virus; this was a higher percentage than in other high-income countries such as the United States and United Kingdom where approximately 60% of the population expressed that they were scared of contracting COVID-19.^21^ Notably, it is also possible that these events may have had an especially damaging impact on some people’s economic situation as the Japanese government announced in April 2020 that the economy is now ‘rapidly deteriorating’ for the first time since 2009.^22^

Hence, the aim of this study was to extend COVID-19-related mental health research beyond China by determining the mental health status of the Japanese population during the pandemic with a particular focus on identifying the most vulnerable groups. In particular, we hypothesized that older adults may be especially at risk for poorer mental health given the higher COVID-19 mortality rates among those aged 70 and above, as well as economically vulnerable individuals due to the possible economic consequences of the COVID-19 pandemic.

## 2. Methods

We administered two rounds of an online survey of the Japanese population between April 16 and April 18, 2020 (1^st^ round) and May 15 and May 17 (2^nd^ round). We utilized a commercial survey company, the Survey Research Center, to send out a set of screening questions to approximately 10,000 respondents from its commercial web panel and then to construct a sample of 1,000 respondents based on their demographic characteristics in each round. A new set of respondents was drawn in the second round. The final sample comprised respondents who were representative of the Japanese general population in terms of the area of their residency, sex, and age distribution. The respondents in the final sample answered online questions about their mental health, personal economic situation, and preventive behavior regarding COVID-19, among others. The final sample size was 2,000.

### 2.1 Ethics statement

This study was approved by the Ethics Committee of Waseda University (approval case number: 2020-050) and Osaka School of International Public Policy, Osaka University. The survey participants were informed of the purpose of the study prior to their participation and had the option to quit the survey at any time. The respondents provided explicit consent that the information they provided could be used for the purpose of this study. The data are completely anonymous.

### 2.2 Measures of mental health status

The Patient Health Questionnaire (PHQ-9)^23^ was used to measure depressive symptoms. The PHQ-9 is a self-report questionnaire that asks about the nine items in the Major Depressive Disorder module in the Diagnostic and Statistical Manual of Mental Disorders. Based on the recommendation in DSM-5, symptoms over the past week were evaluated. The PHQ-9 was translated into Japanese through back translation. Prior studies confirmed the reliability and validity of the Japanese version of this measure.^24,25^ The sensitivity and specificity of the Japanese version of the PHQ-9 were 84% and 95%, respectively.^24^ For each item, responses are rated on a 4-point scale from 0 (“Not at all”) to 3 (“Nearly every day”), with higher scores representing increased levels of depression (total score range of 0–27). For the purpose of this study, we treated a score of 10 or above (“moderate”, “moderately severe”, and “severe” cases) as a case of depressive symptomatology.^26^ The Cronbach’s alpha value for the scale was 0.90.

The 7-item Generalized Anxiety Disorder Scale (GAD-7)^27^ was used to measure anxiety symptoms. The GAD-7 was developed as a self-administered questionnaire for the simple assessment of generalized anxiety disorder. Participants were asked to choose from the following frequency options concerning how often they had been bothered by each of the seven symptoms over the past two weeks: “Not at all”, on “Several days”, on “More than half the days” and “Nearly every day.” The GAD-7 was translated into Japanese through back translation^28^ with prior studies confirming the reliability and validity of the Japanese version.^28,29,30^ The sensitivity and specificity of the Japanese version of the GAD-7 were 88% and 82%, respectively.^28^ For each item, responses are rated on a 4-point scale from 0 to 3, with higher scores representing increased levels of anxiety (total score range 0–21). In the subsequent analysis, we treated a score of 10 or higher (“moderate” and “severe” cases) as a case of anxiety disorder.^31^ The scale had a good degree of internal reliability (Cronbach’s alpha = 0.92).

### 2.3 Demographic information

Information pertaining to basic demographic factors was collected – sex and age. For the age variable, three categories were created; (1) age 18-39 years (young age); (2) age 40-59 years (middle age); (3) 60 years and above (older age). We also included other demographic information on sex and educational attainment, by creating a dummy variable for “male” and “college-educated”. The prefectural area of residency was captured by a dummy variable that took a value of one for individuals who resided in the prefectures that were designated as “special alert areas” (thirteen and eight prefectures at the time of the 1^st^ and 2^nd^ survey, respectively). There were more confirmed cases of COVID-19 in the special alert areas.

### 2.4 Employment status, household income, and household financial situation

To capture the potentially varying effects of the worsening economy and job prospects, we categorized the respondents’ employment status into five mutually exclusive categories: (1) employed as a permanent employee; (2) employed as a short-term contractor, part-time or dispatch worker; (3) self-employed; (4) currently not working but in the labor force; and (5) not in the labor force. Those in the fourth category were either looking for a job, had been laid off, or were taking time off from work (i.e. they were still economically active). The fifth category included homemakers, students, and retired individuals (who were economically inactive). We used the fifth category as the baseline in the subsequent analysis as these individuals are the least likely to be affected by the economic consequences of the pandemic. Treating students (N=21) separately did not change any of the substantive results reported below.

The respondent’s current financial situation was assessed by asking them how they would rate their household’s current financial situation compared to one year ago. Those who answered, “somewhat worse” or “severely worse” were categorized as being “Financially worse off”. This was included in the analysis as a dummy variable.

The household income level of the respondents was also included in the analysis. This was divided into four categories based on annual household income in 2019: (1) less than 4 million yen; (2) equal to or greater than 4 million yen and less than 8 million yen; (3) equal to or greater than 8 million yen; (4) no information. The last category captured respondents who refused to answer or who did not know their annual household income. As this final category included a large number of respondents (N=354, 17.7%), we created a dummy variable that included these respondents and included it in the analysis in order to maximize statistical power. On April 16, 1 JPY=0.009255 USD.

### 2.5 Data Analysis

We first calculated the prevalence of anxiety and depressive symptoms for each of the demographic and economic groups. Then logistic regression models were estimated with either the GAD-7 or PHQ-9 categories as the outcome and all the respondents’ characteristics as the regressors. In each fully adjusted model, we included the respondent’s sex (reference category: female), educational attainment (reference: not college-educated), income level (four categories, reference: 8 million yen and higher), household financial situation (reference: economically not worse off), employment status and type of employment (five categories, reference: not in the labor force), the area of residency (reference: not in the special alert area), and the study period (reference: April 2020). The results are depicted graphically as odds ratios (OR), with the horizontal bars representing 95% confidence intervals (CI). The standard errors were heteroskedasticity-robust, and clustered by prefecture. The analysis was conducted using STATA/MP (version 16, Stata Corporation, College Station, TX). The level of statistical significance was set at p < 0.05 (two-tailed).

## 3. Results

Descriptive statistics of the sample characteristics stratified by the mental health variables are presented in Table 1. Just under half of the respondents were male (49.6%). Almost half (44.7%) of the participants were college-educated, 37.1% were permanent employees, 10.7% were part-time or temporary workers, while 6.9% were not in employment but in the labor force. Fifty-eight percent of the respondents resided in one of the “special alert areas”.

**Table 1.** Descriptive statistics and the prevalence of depressive and anxiety symptoms

| | | % in the sample | % PHQ-9≥10 | % GAD-7≥10 |
| --- | --- | --- | --- | --- |
| All |  |  | 17.3 | 10.9 |
| Age | Young (age 18-39 yrs) | 30.2 | 23.8 | 14.2 |
|  | Middle (age 40-59 yrs) | 36.1 | 20.8 | 14.1 |
|  | Older (age 60 yrs and above) | 33.7 | 7.9 | 4.5 |
| Sex | Female | 50.4 | 17.4 | 10.2 |
|  | Male | 49.6 | 17.3 | 11.6 |
| Education | Without college degree | 55.4 | 18.3 | 11.1 |
|  | With college degree | 44.7 | 16.1 | 10.6 |
| Income | <4 million yen | 32.5 | 22.5 | 13.4 |
| | ≥4 and <8 million yen | 33.6 | 15.6 | 9.4 |
|  | 8 million yen or higher | 16.2 | 13.3 | 9.3 |
|  | No information | 17.7 | 15.0 | 10.7 |
| Household Finance | Unchanged or better off | 79.1 | 13.9 | 8.4 |
|  | Worse off | 21.0 | 30.3 | 20.3 |
| Employment | Permanent Employee | 37.1 | 18.9 | 11.7 |
|  | Part-time, temporary worker | 10.7 | 20.6 | 12.6 |
|  | Self-employed | 4.3 | 9.3 | 4.7 |
|  | Unemployed, laid off, or on leave | 6.9 | 31.2 | 25.4 |
|  | Not in the labor force | 41.0 | 13.7 | 7.9 |
| Area of Residence | Outside special alert area | 42.0 | 17.4 | 11.0 |
|  | In special alert area | 58.0 | 17.3 | 10.9 |

Note: $n=2,000$ . PHQ: the 9-item Patient Health Questionnaire, GAD-7: the 7-item Generalized Anxiety Disorder Scale.

### 3.1 PHQ-9

Among the 2,000 respondents, 17.3% were at or above the cutoff point of 10 for the PHQ-9, indicating the presence of depressive symptoms. The overall mean score on the PHQ-9 was 5.25 (S.D. = 5.62). Based on their PHQ-9 scores, 56.8% of individuals were categorized as having “no” depressive symptoms (PHQ-9 score: 0-4), 25.9% had “mild” symptoms (5-9), 9.5% had “moderate” symptoms (10-14), 5.0% had “moderately severe” symptoms (15-19), while 2.9% had “severe” symptoms (20-27).

### 3.2 GAD-7

As for general anxiety disorder, 218/2000 (10.9%) individuals had a GAD-7 score ≥10. The mean score was 3.73 (S.D. = 4.61). In terms of symptomatology, 68.4% individuals had “none” (GAD-7 score: 0-4), while 20.8% had “mild” (5-9), 6.8% had “moderate” (10-14), and 4.2% had “severe” (15-21) anxiety symptoms.

### 3.3 Logistic Regression analysis

In fully-adjusted models, the odds for at least moderate depressive symptoms were 4.127 (95% CI: 2.683 – 6.348) and 3.385 (95% CI: 2.218 – 5.165) times higher for young and middle-aged respondents, respectively, compared to older respondents (Figure 1). Young (OR: 3.850, 95% CI: 2.542 – 5.831) and middle-aged adults (OR: 3.784, 95% CI: 2.434 – 5.883) also had significantly higher odds for reporting anxiety symptoms compared to their older counterparts (Figure 2).

**Figure 1.**
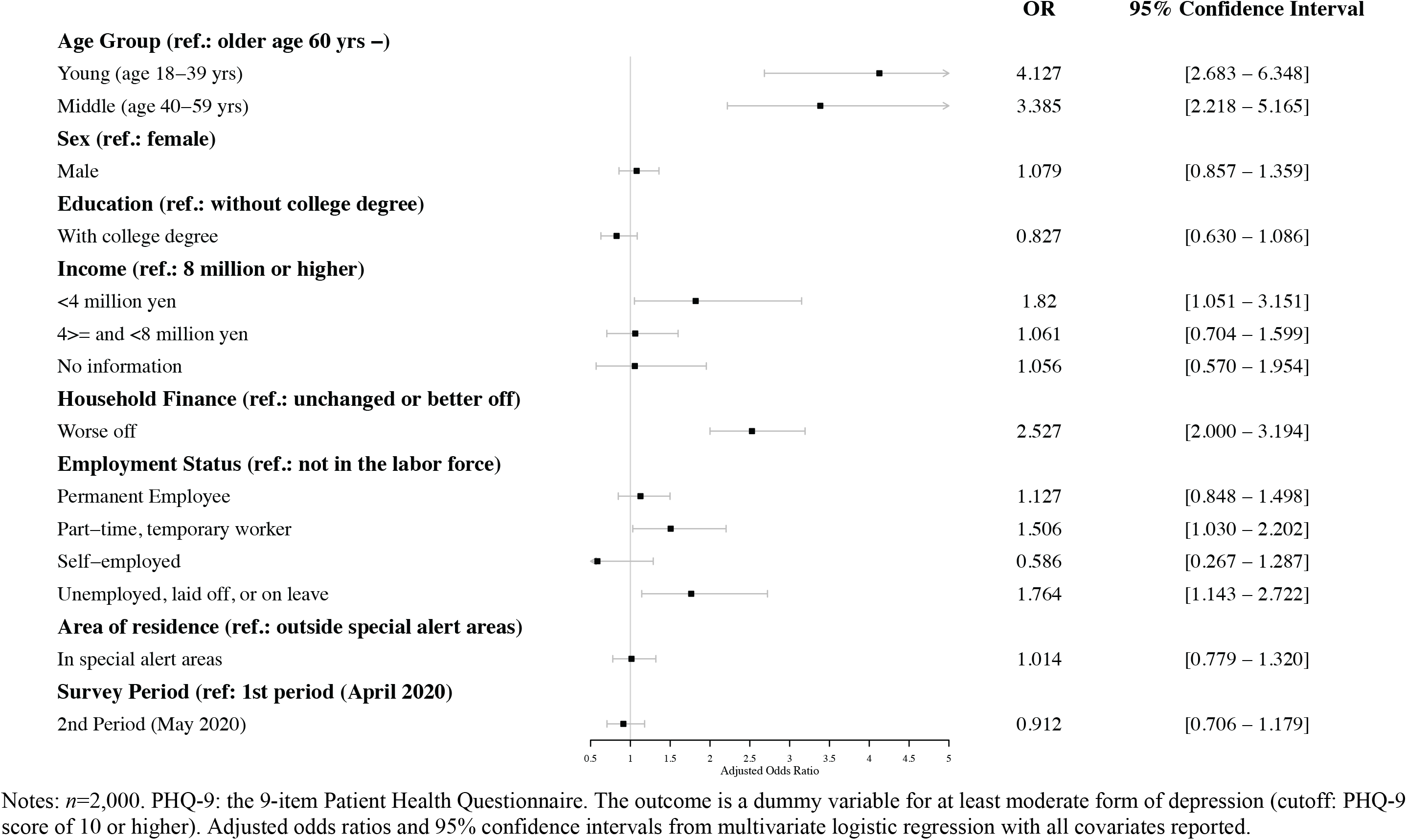
Logistic Regression Results for the Prevalence of Depression Symptoms (PHQ-9>=10)

**Figure 2.**
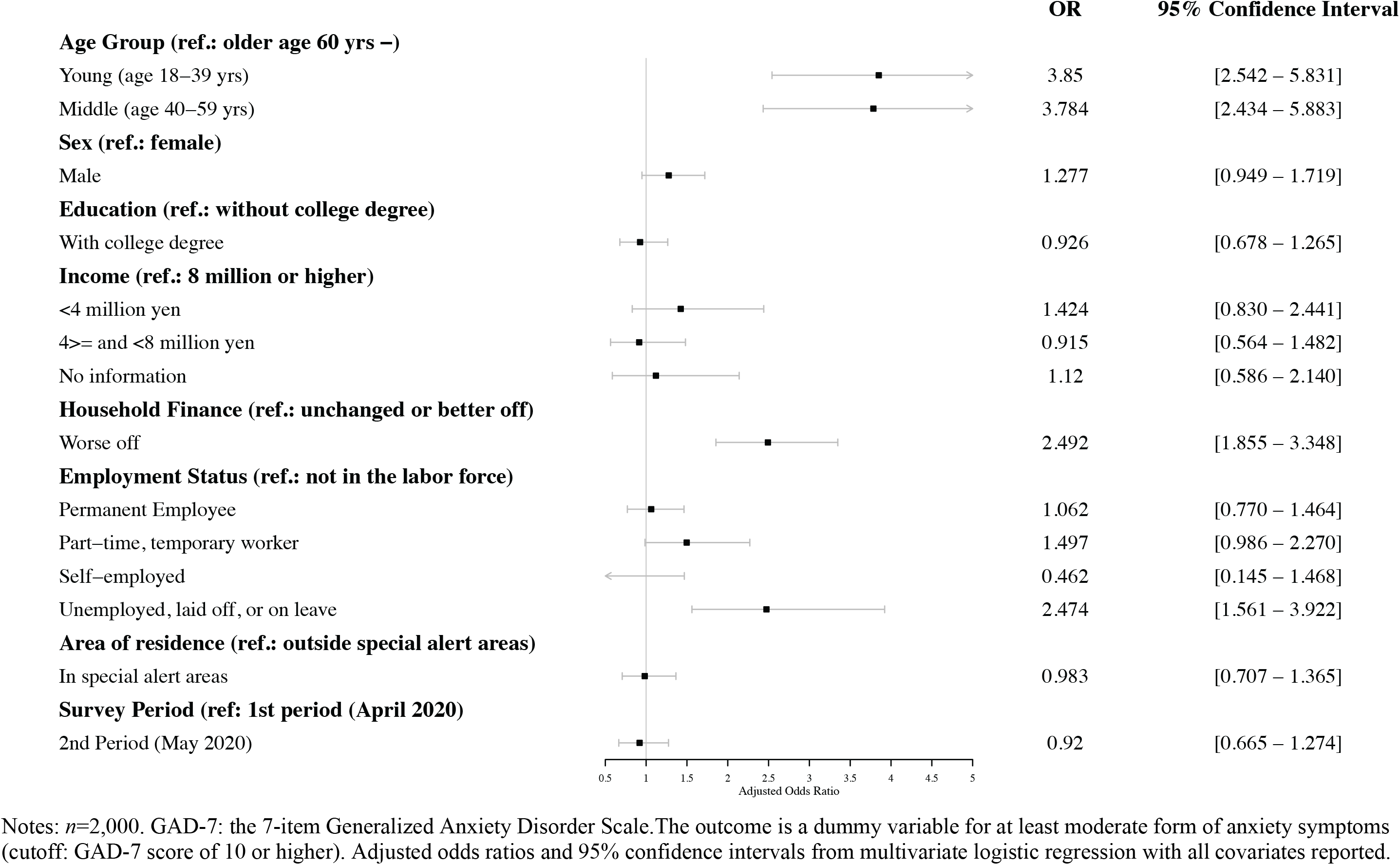
Logistic Regression Results for the Prevalence of Anxiety Symptoms (GAD-7>=10)

Those who were unemployed, had been laid off, or who were taking time off work were 1.764 (95% CI: 1.143 – 2.722) times more likely to have moderate or severe forms of depression compared to those who were not in the labor force, such as retirees and housekeepers. They also had 2.474 (95% CI: 1.561 – 3.922) times higher odds for anxiety symptoms. Individuals with an unstable job (i.e. part-time/temporary workers) had a higher risk of depressive (OR: 1.506, 95% CI: 1.030 – 2.202) and anxiety symptoms (OR: 1.497, 95% CI: 0.986 – 2.270). The odds ratios for permanent workers to experience depression were higher but not statistically different from those not in the labor force.

Respondents who felt that they were financially worse-off compared to a year ago had 2.527 (95% CI: 2.000 – 3.194) and 2.492 (95% CI: 1.855 – 3.348) times higher odds for depressive and anxiety symptoms, respectively, compared to those whose financial situation had not deteriorated. Similarly, the adjusted odds ratio for depressive symptoms for low income individuals was 1.820 (95%: 1.051 – 3.151). Differences in respondents’ sex, and area of residency (whether or not they lived in a special alert area) were not associated with either the prevalence of anxiety or depressive symptoms. Finally, there was no statistically significant difference in the mental health status of respondents between April and May 2020 either in terms of anxiety or depressive symptoms.

## 4. Discussion

This study examined the mental health status of the general population in Japan using information from an online survey of 2000 individuals collected in April and May 2020 that approximated the demographic characteristics of the whole population. To the best of our knowledge, this study constitutes the first investigation of the prevalence of anxiety and depressive symptoms in Japan during the COVID-19 pandemic using a representative sample in terms of age, sex, and regional distribution.

Results indicated that the overall mental health condition of people in Japan may have deteriorated compared to the pre-COVID-19 period. Specifically^32^, as of 2013, 8% of the general population in Japan (N= 3,753) had a PHQ-9 score of 10 or higher. This compares to 17.3% of respondents in the current study. It should be noted that the number of confirmed cases and deaths from COVID-19 is still relatively low in Japan at the time of writing this manuscript, compared to other affected countries; as of April 27, 2020, there were 360 deaths attributed to COVID-19, which corresponds to 2.8 per one million persons. For comparison, the number of confirmed deaths per one million population in the United States was 164.3.^33^

The findings of our study suggest that the mental health condition of some segments of the Japanese population may be particularly vulnerable during the ongoing COVID-19 crisis. In particular, individuals who were not currently working were more likely to report a high level of anxiety and depressive symptoms. Our results also indicate that the form of employment might also be important for mental health; individuals who are employed as part-time or temporary contract-based workers were more likely to suffer from depressive symptoms compared to the reference group. The finding that non-/precarious employment may be linked to poorer mental health accords with the finding of a recent study of 369 adults in 64 cities in China where stopping work during the COVID-19 crisis was associated with worse mental health.^34^

These results are consistent with the notion that the effects of a faltering economy and reduction in business activities caused by COVID-19 are first and foremost likely to detrimentally affect workers without stable employment. When various non-essential businesses were closed following the declaration of a state of emergency, many part-time workers immediately lost their source of income. Moreover, for those who were seeking employment, their prospects of finding a job have now greatly diminished as the number of job listings has declined.^22^ Thus, it is possible that the socioeconomic stresses and strains that are emerging as a result of the ongoing pandemic might have had an especially hard impact on these precarious workers and jobseekers in terms of poorer mental health.

Similarly, individuals who felt that their financial position deteriorated in the past year had greater odds for depression and anxiety disorder. As we controlled for the effect of income level on mental health in the analyses, it suggests that these results cannot be entirely explained by the differences in income levels. Although it is uncertain what underlies this specific finding, taken together with the results relating to employment status, it suggests that individuals who are economically vulnerable might be especially susceptible to experiencing poorer mental health during the COVID-19 pandemic, especially if the economic effects of the crisis are protracted.

It is important to note however, that the mental health condition of other employment status groups may have also deteriorated compared to the pre-COVID-19 period. Specifically, earlier research undertaken in Japan in 2013, showed that 5.84% of full-time workers in the general population could be classified as being depressed based on a PHQ9 cutoff score ≥ 10.^32^ In contrast, 18.9% of permanent employees in our study met the same criterion (Table 1). In addition, the prevalence of depression among homemakers, retirees, and students (the reference group in our study) was reported to be 8.25% in 2013 whereas 13.7% reported depressive symptoms in the current study. Thus, there was a 13.06 point difference for full-time workers and a 5.45 point difference for those not in the labor force. This suggests that the mental health conditions of all workers (both economically active and inactive) should be closely monitored during and following the current crisis as the negative economic shocks associated with COVID-19 might affect all employment status categories.

Contrary to our hypothesis, we found strong evidence that the mental health of young and middle-aged individuals may be significantly worse than that of older individuals during the pandemic. To date, studies from China have produced mixed findings concerning the mental health of different age groups during the COVID-19 crisis.^14,35,36^ It is not immediately obvious as to why the mental health of the elderly population is significantly better than that of younger individuals. Similar to elsewhere in the world, they are by far the highest-risk group for death and complications by COVID-19. For example, out of 348 deaths due to COVID-19 in Japan as of April 26, 2020, 228 (66%) were among persons aged 60 and above.^37^ In addition, in the abovementioned Japanese study from 2013,^32^ the elderly population (aged ≥ 65) had a higher prevalence of depression compared to those who were young and middle-aged.

Although more work needs to be done to understand the impact of COVID-19 on different age groups, it can be speculated that the crisis might be presenting a much greater range of difficulties for the working-age population, rather than older age groups. For example, besides financial worries, it is also possible that COVID-19 may be currently giving rise to other stressors in younger age groups that might also impact their mental health – such as the need for parents to both telework from home while at the same time homeschool their children. Support for this notion of a greater number/different range of stressors in younger age groups comes from a recent study from China which linked worse mental health in those aged 12-21 years with education difficulties arising from both school closures and exam uncertainties.^14^ Alternatively, it is possible that other factors might be important in this context. An earlier study found, for example, that older adults were more psychologically resilient than their younger counterparts^38^ which might be important when it comes to dealing with the sources of stress associated with the COVID-19 pandemic.

This study has several strengths. It is the first systematic study to examine the mental health status of the general population in Japan during the COVID-19 pandemic. While doing this, we were able to draw on data from a large population-based sample. Moreover, we were also able to examine different mental health outcomes. Despite this, our study also has several limitations that should be mentioned. First, our survey was undertaken among respondents from a commercial web panel, and thus did not guarantee that the sample would be perfectly representative of the entire population. Although being able to conduct the survey while using random sampling methodology would have been preferable, it was not appropriate in this particular instance as it would not have allowed us to collect data in a timely manner – which was essential given the urgent need to understand how COVID-19 is affecting population well-being. Second, our study is cross-sectional, and thus cannot establish a causal relationship between the pandemic and mental health conditions. In particular, those who are in an economically weak position are likely to be vulnerable to poorer mental health even during non-crisis times^39^, and our study could not prove that their mental health was poorer because of any major economic upheaval associated with COVID-19. However, the main purpose of this study was not to establish causality; its focus was rather on identifying the most vulnerable groups during the initial stage of the pandemic in Japan by undertaking the first systematic research on the mental health of the Japanese population during this period. We are planning to conduct a series of similar surveys in the following months that should further elucidate the effects of COVID-19 on the population’s mental health. Third, our analysis did not consider the potential effects of marital status which might have been important for mental health.

Despite these limitations, our study contributes to the literature by identifying the most vulnerable population groups during the COVID-19 crisis through the first systematic survey of the mental health of the Japanese general population. Our results highlight that monitoring the mental health of younger and economically vulnerable individuals may be especially important moving forward. In addition, they also indicate that the general public’s mental health during the pandemic might not only be affected by the direct health consequences of COVID-19, but also by the economic ramifications of the pandemic.

The results of the present study also have important policy implications. Given the potentially devastating impact of the pandemic on the economy, not just in Japan, but worldwide, policymakers should provide financial aid to those affected by the economic consequences of the pandemic. In connection with this, the Japanese government had initially planned to distribute a cash benefit of 300,000 yen to each household provided that the head of the household had lost a significant portion of their income, but then changed the planned policy in mid-April so that all residents in Japan would receive 100,000 yen per person.^40^ While funds are expected to reach residents faster under the current universal policy, many small-sized households that have been economically affected will receive less money than they would have under the previous policy. This suggests that the government should employ both universal and targeted policies so that the needs of the most economically vulnerable are met during this unprecedented crisis.

## Data Availability

Due to the sensitive nature of the questions asked in this study, survey respondents were assured that individual raw data would not be shared.

## Acknowledgements

This work was financially supported by JSPS Grants-in-Aid for Scientific Research Grant Number 20H01584. The funders had no role in study design, data collection and analysis, decision to publish, or preparation of the manuscript.

---

**Supplementary Table 1:** PHQ-9 Scores by Subgroup

|  |  | count | mean | std | min | 25% | 50% | 75% | max |
| --- | --- | --- | --- | --- | --- | --- | --- | --- | --- |
| All | All | 2000 | 5.250 | 5.616 | 0 | 1 | 4 | 8 | 27 |
| Age groups (3 groups) | Young (age 18-39 yrs) | 604 | 6.364 | 6.172 | 0 | 1 | 5 | 9 | 27 |
|  | Middle (age 40-59 yrs) | 722 | 5.852 | 5.930 | 0 | 1 | 4 | 9 | 27 |
|  | Older (age 60 yrs -) | 674 | 3.607 | 4.204 | 0 | 0 | 2 | 5 | 27 |
| Age groups (6 groups) | 18-29 yrs | 304 | 6.885 | 6.302 | 0 | 2 | 5 | 10 | 27 |
|  | 30-39 yrs | 300 | 5.837 | 6.001 | 0 | 1 | 4 | 9 | 27 |
|  | 40-49 yrs | 380 | 6.321 | 6.222 | 0 | 1 | 5 | 9 | 27 |
|  | 50-59 yrs | 342 | 5.330 | 5.550 | 0 | 1 | 4 | 8 | 27 |
|  | 60-69 yrs | 354 | 4.020 | 4.773 | 0 | 0 | 3 | 6 | 27 |
|  | 70 yrs- | 320 | 3.150 | 3.417 | 0 | 0 | 2 | 5 | 18 |
| Sex | Female | 1008 | 5.288 | 5.441 | 0 | 1 | 4 | 8 | 27 |
|  | Male | 992 | 5.212 | 5.791 | 0 | 1 | 3 | 8 | 27 |
| Education | Without college degree | 1107 | 5.353 | 5.757 | 0 | 1 | 4 | 8 | 27 |
|  | With college degree | 893 | 5.122 | 5.437 | 0 | 1 | 4 | 8 | 27 |
| Household income | <4 million yen | 650 | 6.165 | 6.157 | 0 | 1 | 4 | 9 | 27 |
|  | 4>= and <8 million yen | 672 | 4.714 | 5.049 | 0 | 1 | 3 | 7 | 27 |
|  | 8 million or higher | 324 | 4.676 | 5.216 | 0 | 1 | 3 | 7 | 27 |
|  | No information | 354 | 5.113 | 5.757 | 0 | 1 | 3 | 7 | 27 |
| Household Finance | Unchanged or better off | 1581 | 4.672 | 5.205 | 0 | 1 | 3 | 7 | 27 |
|  | Worse off | 419 | 7.432 | 6.511 | 0 | 2 | 6 | 11 | 27 |
| Employment Status | Permanent Employee | 742 | 5.453 | 5.670 | 0 | 1 | 4 | 8 | 27 |
|  | Part-time, temporary worker | 214 | 5.374 | 5.184 | 0 | 1 | 5 | 9 | 26 |
|  | Self-employed | 86 | 3.814 | 4.677 | 0 | 0 | 3 | 5 | 27 |
|  | Unemployed, laid off, or on leave | 138 | 8.058 | 7.333 | 0 | 2 | 6 | 11.75 | 27 |
|  | Not in the labor force | 820 | 4.712 | 5.269 | 0 | 1 | 3 | 7 | 27 |
| Area of residence | Outside special alert areas | 840 | 5.269 | 5.716 | 0 | 1 | 4 | 8 | 27 |
|  | In special alert areas | 1160 | 5.236 | 5.545 | 0 | 1 | 4 | 8 | 27 |

Note: PHQ-9 is the 9-item Patient Health Questionnaire (range 0-27).

**Supplementary Table 2:**
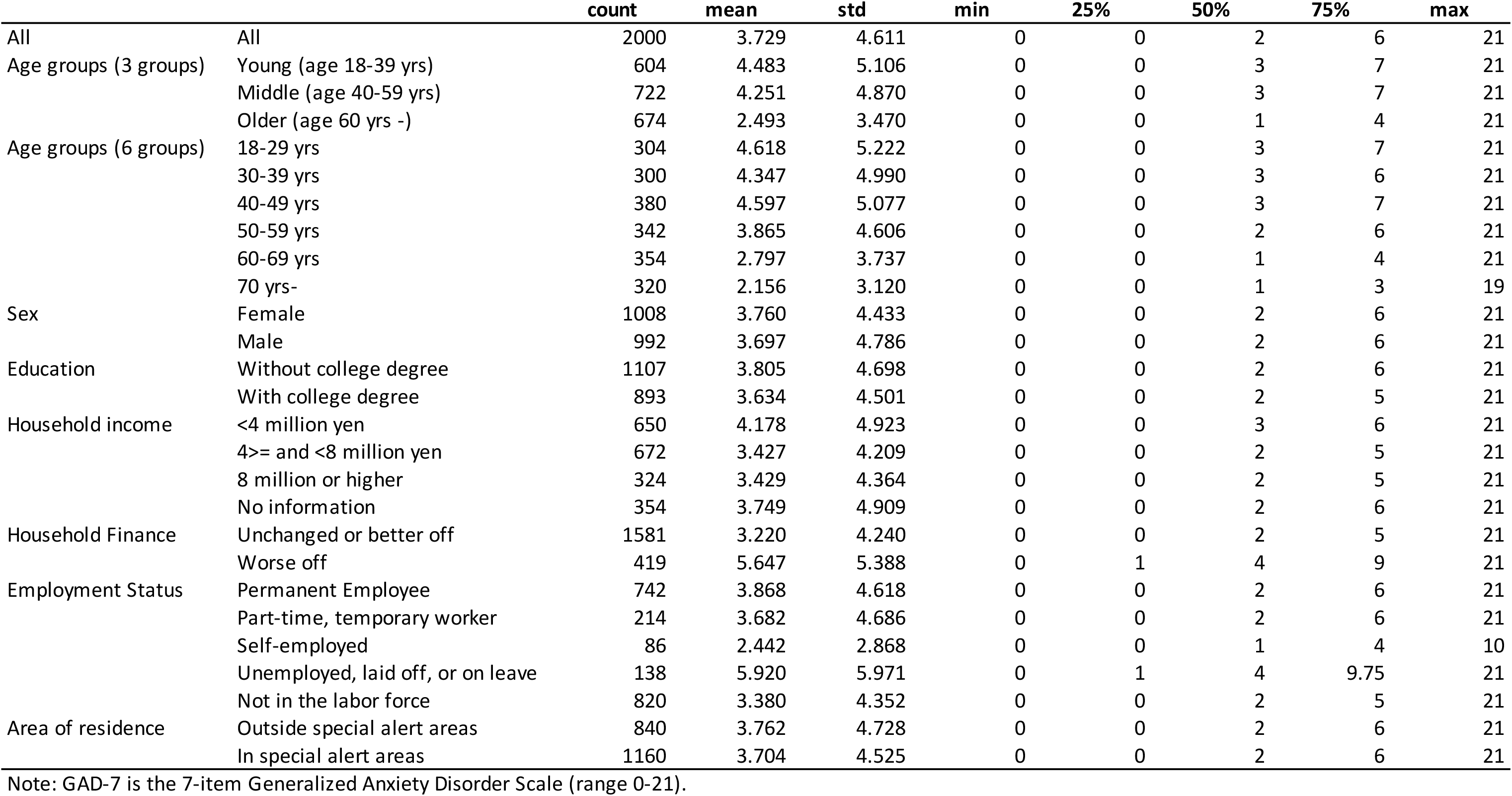
GAD-7 Scores by Subgroup

**Supplementary Table 3:** PHQ-9 Categories by Subgroup

|  |  | n |  |  |  |  |  | % |  |  |  |  |
| --- | --- | --- | --- | --- | --- | --- | --- | --- | --- | --- | --- | --- |
|  |  | count | none | mild | moderate | moderately severe | severe | none | mild | moderate | moderately severe | severe |
| All | All | 2000 | 1136 | 517 | 190 | 99 | 58 | 56.8 | 25.9 | 9.5 | 5.0 | 2.9 |
| Age groups (3 groups) | Young (age 18-39 yrs) | 604 | 300 | 160 | 73 | 43 | 28 | 49.7 | 26.5 | 12.1 | 7.1 | 4.6 |
|  | Middle (age 40-59 yrs) | 722 | 374 | 198 | 81 | 44 | 25 | 51.8 | 27.4 | 11.2 | 6.1 | 3.5 |
|  | Older (age 60 yrs -) | 674 | 462 | 159 | 36 | 12 | 5 | 68.5 | 23.6 | 5.3 | 1.8 | 0.7 |
| Age groups (6 groups) | 18-29 yrs | 304 | 137 | 89 | 40 | 20 | 18 | 45.1 | 29.3 | 13.2 | 6.6 | 5.9 |
|  | 30-39 yrs | 300 | 163 | 71 | 33 | 23 | 10 | 54.3 | 23.7 | 11.0 | 7.7 | 3.3 |
|  | 40-49 yrs | 380 | 185 | 102 | 51 | 26 | 16 | 48.7 | 26.8 | 13.4 | 6.8 | 4.2 |
|  | 50-59 yrs | 342 | 189 | 96 | 30 | 18 | 9 | 55.3 | 28.1 | 8.8 | 5.3 | 2.6 |
|  | 60-69 yrs | 354 | 229 | 86 | 28 | 6 | 5 | 64.7 | 24.3 | 7.9 | 1.7 | 1.4 |
|  | 70 yrs- | 320 | 233 | 73 | 8 | 6 | 0 | 72.8 | 22.8 | 2.5 | 1.9 | 0.0 |
| Sex | Female | 1008 | 565 | 268 | 99 | 50 | 26 | 56.1 | 26.6 | 9.8 | 5.0 | 2.6 |
|  | Male | 992 | 571 | 249 | 91 | 49 | 32 | 57.6 | 25.1 | 9.2 | 4.9 | 3.2 |
| Education | Without college degree | 1107 | 627 | 277 | 113 | 53 | 37 | 56.6 | 25.0 | 10.2 | 4.8 | 3.3 |
|  | With college degree | 893 | 509 | 240 | 77 | 46 | 21 | 57.0 | 26.9 | 8.6 | 5.2 | 2.4 |
| Household income | <4 million yen | 650 | 333 | 171 | 79 | 39 | 28 | 51.2 | 26.3 | 12.2 | 6.0 | 4.3 |
|  | 4>= and <8 million yen | 672 | 409 | 158 | 65 | 29 | 11 | 60.9 | 23.5 | 9.7 | 4.3 | 1.6 |
|  | 8 million or higher | 324 | 191 | 90 | 20 | 17 | 6 | 59.0 | 27.8 | 6.2 | 5.2 | 1.9 |
|  | No information | 354 | 203 | 98 | 26 | 14 | 13 | 57.3 | 27.7 | 7.3 | 4.0 | 3.7 |
| Household Finance | Unchanged or better off | 1581 | 962 | 399 | 126 | 61 | 33 | 60.8 | 25.2 | 8.0 | 3.9 | 2.1 |
|  | Worse off | 419 | 174 | 118 | 64 | 38 | 25 | 41.5 | 28.2 | 15.3 | 9.1 | 6.0 |
| Employment Status | Permanent Employee | 742 | 415 | 187 | 73 | 49 | 18 | 55.9 | 25.2 | 9.8 | 6.6 | 2.4 |
|  | Part-time, temporary worker | 214 | 106 | 64 | 31 | 11 | 2 | 49.5 | 29.9 | 14.5 | 5.1 | 0.9 |
|  | Self-employed | 86 | 60 | 18 | 6 | 0 | 2 | 69.8 | 20.9 | 7.0 | 0.0 | 2.3 |
|  | Unemployed, laid off, or on leave | 138 | 56 | 39 | 17 | 11 | 15 | 40.6 | 28.3 | 12.3 | 8.0 | 10.9 |
|  | Not in the labor force | 820 | 499 | 209 | 63 | 28 | 21 | 60.9 | 25.5 | 7.7 | 3.4 | 2.6 |
| Area of residence | Outside special alert areas | 840 | 474 | 220 | 79 | 38 | 29 | 56.4 | 26.2 | 9.4 | 4.5 | 3.5 |
|  | In special alert areas | 1160 | 662 | 297 | 111 | 61 | 29 | 57.1 | 25.6 | 9.6 | 5.3 | 2.5 |

Note: "none" refers to PHQ-9 score of 0-4; "mild": 5-9; "moderate": 10-14; "moderately severe": 15-19; "severe": 20-27.

**Supplementary Table 4:** GAD-7 Categories by Subgroup

|  |  | count | n |  |  |  | % |  |  |  |
| --- | --- | --- | --- | --- | --- | --- | --- | --- | --- | --- |
|  |  |  | none | mild | moderate | severe | none | mild | moderate | severe |
| All | All | 2000 | 1367 | 415 | 135 | 83 | 68.4 | 20.8 | 6.8 | 4.2 |
| Age groups (3 groups) | Young (age 18-39 yrs) | 604 | 376 | 142 | 49 | 37 | 62.3 | 23.5 | 8.1 | 6.1 |
|  | Middle (age 40-59 yrs) | 722 | 462 | 158 | 63 | 39 | 64.0 | 21.9 | 8.7 | 5.4 |
|  | Older (age 60 yrs -) | 674 | 529 | 115 | 23 | 7 | 78.5 | 17.1 | 3.4 | 1.0 |
| Age groups (6 groups) | 18-29 yrs | 304 | 190 | 67 | 28 | 19 | 62.5 | 22.0 | 9.2 | 6.3 |
|  | 30-39 yrs | 300 | 186 | 75 | 21 | 18 | 62.0 | 25.0 | 7.0 | 6.0 |
|  | 40-49 yrs | 380 | 232 | 86 | 38 | 24 | 61.1 | 22.6 | 10.0 | 6.3 |
|  | 50-59 yrs | 342 | 230 | 72 | 25 | 15 | 67.3 | 21.1 | 7.3 | 4.4 |
|  | 60-69 yrs | 354 | 270 | 64 | 15 | 5 | 76.3 | 18.1 | 4.2 | 1.4 |
|  | 70 yrs- | 320 | 259 | 51 | 8 | 2 | 80.9 | 15.9 | 2.5 | 0.6 |
| Sex | Female | 1008 | 683 | 222 | 66 | 37 | 67.8 | 22.0 | 6.5 | 3.7 |
|  | Male | 992 | 684 | 193 | 69 | 46 | 69.0 | 19.5 | 7.0 | 4.6 |
| Education | Without college degree | 1107 | 741 | 243 | 73 | 50 | 66.9 | 22.0 | 6.6 | 4.5 |
|  | With college degree | 893 | 626 | 172 | 62 | 33 | 70.1 | 19.3 | 6.9 | 3.7 |
| Household income | <4 million yen | 650 | 414 | 149 | 54 | 33 | 63.7 | 22.9 | 8.3 | 5.1 |
|  | 4>= and <8 million yen | 672 | 473 | 136 | 42 | 21 | 70.4 | 20.2 | 6.3 | 3.1 |
|  | 8 million or higher | 324 | 231 | 63 | 20 | 10 | 71.3 | 19.4 | 6.2 | 3.1 |
|  | No information | 354 | 249 | 67 | 19 | 19 | 70.3 | 18.9 | 5.4 | 5.4 |
| Household Finance | Unchanged or better off | 1581 | 1148 | 300 | 86 | 47 | 72.6 | 19.0 | 5.4 | 3.0 |
|  | Worse off | 419 | 219 | 115 | 49 | 36 | 52.3 | 27.4 | 11.7 | 8.6 |
| Employment Status | Permanent Employee | 742 | 496 | 159 | 59 | 28 | 66.8 | 21.4 | 8.0 | 3.8 |
|  | Part-time, temporary worker | 214 | 148 | 39 | 19 | 8 | 69.2 | 18.2 | 8.9 | 3.7 |
|  | Self-employed | 86 | 67 | 15 | 4 | 0 | 77.9 | 17.4 | 4.7 | 0.0 |
|  | Unemployed, laid off, or on leave | 138 | 74 | 29 | 20 | 15 | 53.6 | 21.0 | 14.5 | 10.9 |
|  | Not in the labor force | 820 | 582 | 173 | 33 | 32 | 71.0 | 21.1 | 4.0 | 3.9 |
| Area of residence | Outside special alert areas | 840 | 574 | 174 | 50 | 42 | 68.3 | 20.7 | 6.0 | 5.0 |
|  | In special alert areas | 1160 | 793 | 241 | 85 | 41 | 68.4 | 20.8 | 7.3 | 3.5 |

Note: "none" refers to GAD-7 score of 0-4; "mild": 5-9; "moderate": 10-14; "severe": 15-21.

**Supplementary Figure 1:**
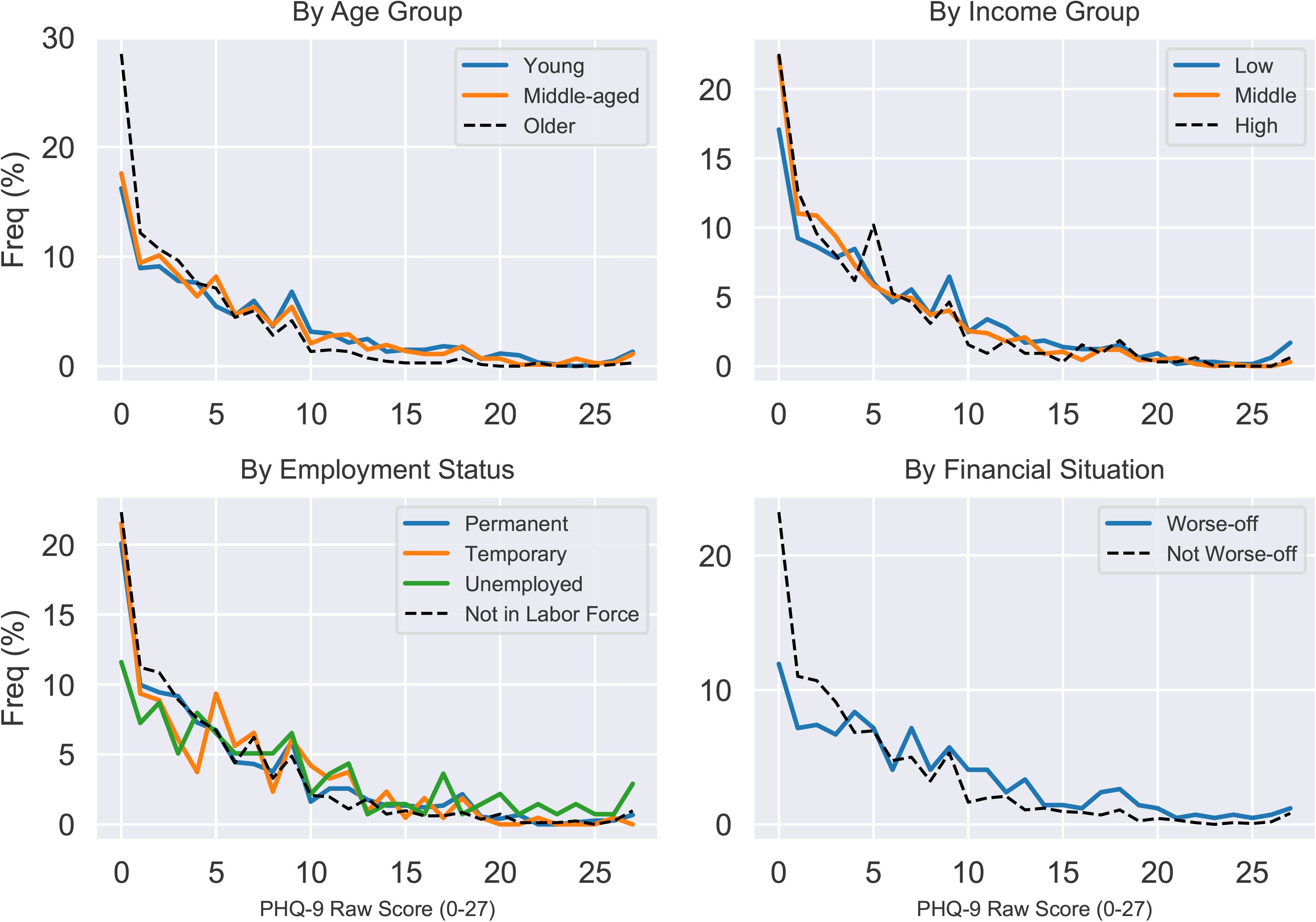
PHQ-9 Raw Scores by Selected Subgroup

**Supplementary Figure 2:**
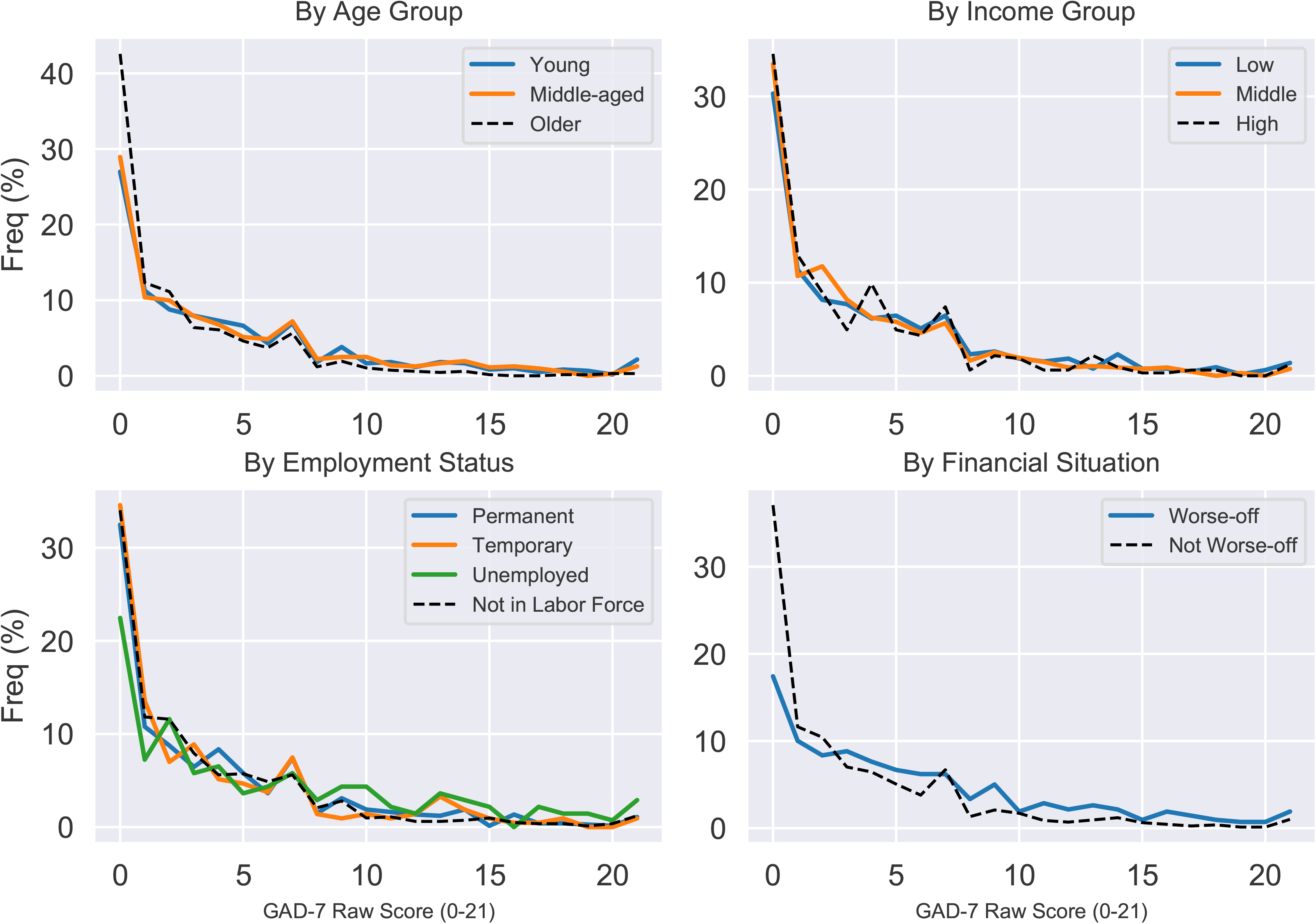
GAD-7 Raw Scores by Selected Subgroup

